# PERADS.net: Automated PE-RADS Grading with Named Anatomic Localization and Right-to-Left Ventricular Ratio Measurement on CT Pulmonary Angiography

**DOI:** 10.64898/2026.09.15.26363142

**Authors:** Ezio Lanza, Federica Catapano, Costanza Lisi, Federico D’Orazio, Riccardo Levi, Andrea Laghi

## Abstract

**Purpose:** To develop an automated pipeline (PERADS.net) that segments acute pulmonary embolism on CT pulmonary angiography, assigns a Pulmonary Embolism Reporting and Data System (PE-RADS) grade with named anatomic localization, and measures the right-to-left ventricular (RV/LV) diameter ratio, and to assess radiologist agreement.

**Materials and Methods:** In this retrospective study, 120 CT pulmonary angiograms (70 peripheral, 35 central, 15 negative) from the public RSNA Pulmonary Embolism CT dataset were analyzed. A two-channel five-fold nnU-Net ensemble segmented the embolus; the pulmonary arterial tree was reduced to a branching graph, and the grade was set by the most proximal level with at least 1% of embolic volume.

Three radiologists, blinded to the algorithm-assigned grade, independently assigned grades and rated RV/LV plausibility. Agreement was assessed with percent agreement and Cohen or Fleiss kappa on five-grade and grouped scales (grade 0 versus 1-2 versus 3-4). The automated ratio was compared with the dataset’s binary RV/LV label in 2119 examinations.

**Results:** Agreement between the algorithm and three-radiologist consensus (n = 119) was 60.5% (kappa, 0.38) for individual grades and 90.8% (kappa, 0.74; 95% CI: 0.59, 0.87) for the grouped scale; 34 of 47 discordant examinations fell within grades 3-4. As a fourth reader, the algorithm matched grouped-scale interobserver agreement (mean kappa, 0.62 versus 0.67). Radiologists showed no agreement rating RV/LV plausibility as favorable or incorrect (Fleiss kappa, −0.00), whereas the automated ratio agreed with the external label (kappa, 0.49), overestimating strain 3.1:1.

**Conclusion:** Automated PE-RADS grading agreed with radiologist consensus comparably to a fourth reader on the grouped scale.

**Summary Statement:** An automated pipeline assigned PE-RADS grades agreeing with three-radiologist consensus at a level approaching interobserver agreement on the clinically grouped scale, while automated RV/LV measurement showed no reader agreement on plausibility.

**Key Points:**

- In 120 CT pulmonary angiograms, agreement between automated and consensus PE-RADS grading was 90.8% (kappa, 0.74; 95% CI: 0.59, 0.87) on the clinically grouped scale (grade 0 versus 1-2 versus 3-4) and 60.5% (kappa, 0.38; 95% CI: 0.24, 0.51) on the five-grade scale.
- Treated as a fourth reader, the algorithm reached grouped-scale agreement with individual radiologists (mean kappa, 0.62) comparable to that observed among radiologists themselves (mean kappa, 0.67).
- Automated right-to-left ventricular ratio agreed moderately with an independent binary label in 2119 examinations (kappa, 0.49; 95% CI: 0.45, 0.52), whereas three radiologists showed no agreement when rating the same measurement as favorable or incorrect (Fleiss kappa, −0.00; 95% CI: −0.09, 0.09).

## Introduction

Acute pulmonary embolism (PE) is common and potentially fatal, and risk-adapted management depends on the extent and central location of clot and on signs of right ventricular strain (1,2). To reduce variability in how these findings are communicated, the Pulmonary Embolism Reporting and Data System (PE-RADS) was introduced in 2026 as a structured lexicon assigning a grade based on the most proximal arterial level involved (3). Adoption in routine practice depends on whether grading can be produced consistently and without adding reporting time.

Deep learning applied to CT pulmonary angiography (CTPA) has focused on detection and triage rather than structured severity reporting (4-7). Available algorithms report the presence of embolism, and some estimate clot burden or right-to-left ventricular (RV/LV) diameter ratio, but a pipeline that segments the embolus, names the most proximal structure, assigns a PE-RADS grade, and measures the RV/LV ratio has not been described.

This study reports the technical development of such a pipeline, PERADS.net, and a preliminary assessment of its agreement with radiologist interpretation. The name denotes the software described here and implies no endorsement by the PE-RADS committee. Because no independent reference standard for PE-RADS grade exists, the algorithm was evaluated in two complementary ways: as a fourth independent reader alongside three radiologists, and against the majority consensus of those radiologists. Agreement, not diagnostic accuracy, is therefore the outcome throughout.

## Materials and Methods

### Study design and dataset

This retrospective study drew on two distinct datasets. The segmentation network was developed on an institutional cohort, whereas all analyses reported here used the publicly available RSNA Pulmonary Embolism CT dataset (8,9), comprising de-identified CTPA examinations annotated at examination and slice level by a multi-institutional panel.

Because the evaluation data are public and fully de-identified, their secondary analysis required no ethics committee review. The development cohort was collected under an institutional policy whereby retrospective studies not affecting clinical care proceed without ethics submission and with waiver of written informed consent. Reporting followed the Checklist for Artificial Intelligence in Medical Imaging (10).

From the public dataset, 120 examinations were drawn by random sampling within severity strata: 70 (58%) with peripheral PE, 35 (29%) with central PE, and 15 (13%) without PE. All were processed from raw, uncropped image data. Patient age and sex were unavailable because the dataset is de-identified.

### Pipeline description

Anatomic reference structures were segmented on uncropped chest CT with TotalSegmentator (11,12), using the pulmonary vasculature, cardiac chamber, and pulmonary lobe tasks. A thoracic bounding box encompassing airways, pulmonary arteries, and pulmonary veins cropped the volume for inference (Fig 1).

**Figure 1.**
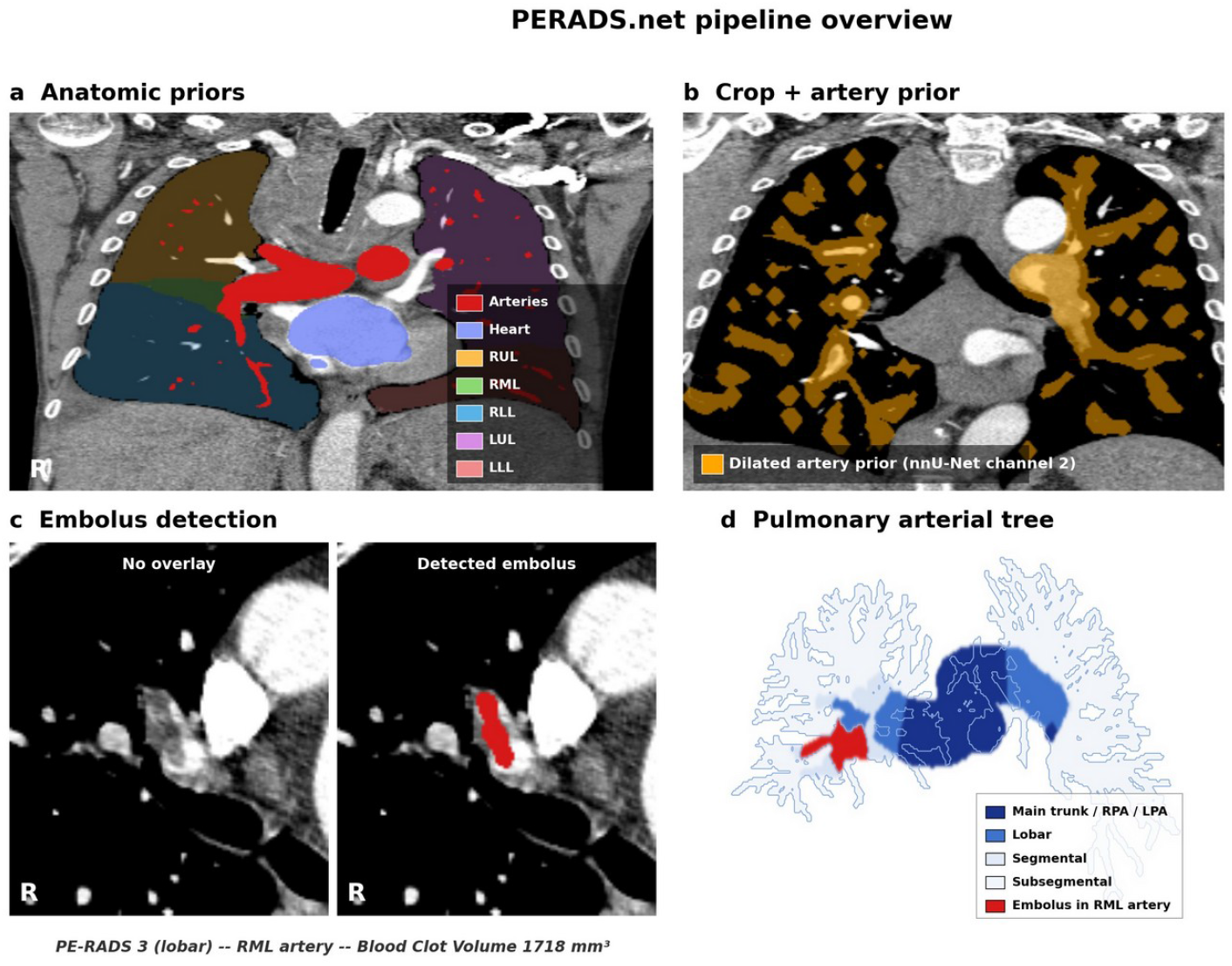
PERADS.net pipeline overview. (a) Anatomic priors derived from TotalSegmentator on the raw, uncropped CT: pulmonary arteries (red), heart (blue), and the five pulmonary lobes (right upper, right middle, right lower, left upper, left lower). (b) Chest CT cropped to the thoracic cavity, with the dilated pulmonary artery prior (orange) supplied as the second input channel to the nnU-Net embolus segmentation model. (c) Axial CT without (left) and with (right) the detected embolus overlaid in red in the right middle lobe artery. (d) Named pulmonary arterial tree, color coded by anatomic level (main trunk and right or left pulmonary artery, lobar, segmental, subsegmental), with the detected embolus (red) localized to the right middle lobe artery. Example case: AI-PE-RADS grade 3 (lobar), embolic volume 1718 mm3; consensus grade 3.

Embolus segmentation used a two-channel three-dimensional full-resolution nnU-Net (13) deployed as a five-fold ensemble. The first channel was the cropped CT volume and the second a dilated pulmonary arterial mask, an anatomic prior. Predictions were realigned to full-resolution space by translation from the affine matrices, then intersected with the arterial mask, discarding extra-arterial voxels. Examinations with less than 70 mm3 of embolic volume were assigned grade 0.

The pulmonary arterial mask was reduced to a branching graph by three-dimensional skeletonization, rooted at the main pulmonary trunk. Branches were assigned to lobes by overlap with lobar masks, with compensatory dilation near the hilum and comparison of caliber between sibling branches; lobar assignment remained revisable. Each branch was labeled as main trunk or right or left pulmonary artery, lobar, segmental, or subsegmental. The grade assigned by PERADS.net, hereafter AI-PE-RADS, corresponded to the most proximal level accounting for at least 1% of embolic volume, and the involved structures were reported by name. AI-PE-RADS denotes pipeline output on the PE-RADS scale, not a separate system.

The RV/LV diameter ratio was derived from a four-chamber method. An anatomic four-chamber plane was computed from cardiac chamber centroids, the left ventricular apex was identified, and ventricular diameters were measured along a line parallel to the chamber base at 25% of the base-to-apex height, taken as separate contiguous intersections of the respective chamber masks (Fig 5). Median processing time was 223 seconds per examination (interquartile range, 193-261 seconds).

The segmentation network was trained on a stratified subset of 437 CTPA examinations, with manually refined embolus contours, from the same institutional dataset described in reference 14.

### Reader study

Three radiologists with 19, 12, and 7 years of post-training experience independently assigned a PE-RADS grade to each examination while viewing the algorithm-generated segmentation overlay, blinded to the algorithm-assigned grade. Case order was randomized at each session. Readers additionally rated four-chamber RV/LV segmentation plausibility as acceptable without reservation, acceptable, or incorrect, and could flag any examination in which embolus segmentation appeared incomplete. Readers rated anatomic plausibility, not clinical validity of the ratio. Because both acceptable categories reflect a favorable judgment, plausibility was analyzed primarily as favorable versus incorrect, with the three-level scale as secondary detail.

### External comparison of the RV/LV ratio

Because the reader study addressed only whether the measurement was anatomically plausible, the ratio was also compared with an independent label. The public dataset provides a binary indication of whether the RV/LV ratio is at least 1, assigned only to examinations with PE present, giving a maximum of 2211 comparable examinations. The pipeline was applied to the entire dataset separately, and its ratio, dichotomized at 1.0, was compared with this label as external corroboration, not a reference standard.

## Statistical analysis

Two parallel analyses were performed, each on the five-grade PE-RADS scale and on a grouped scale (grade 0 versus 1-2 versus 3-4), grouped because these bands track PE-RADS management pathways: none, anticoagulation, or reperfusion consideration (3). First, the algorithm was treated as a fourth independent reader, and pairwise percent agreement and Cohen kappa (15) were computed for all rater pairs. Second, the algorithm was compared with the majority-vote consensus of the three radiologists; one examination without a majority grade was excluded from that comparison. Simultaneous agreement across three or four raters was quantified with Fleiss kappa. Confidence intervals were obtained by bootstrap resampling with 5000 replicates, and kappa values were interpreted according to established categories (16). Analyses were performed in Python 3.11 with scikit-learn 1.8 (Cohen kappa), statsmodels 0.15 (Fleiss kappa), and NumPy 2.4 (bootstrap resampling).

## Results

### Study sample and algorithm output

All 120 examinations were processed successfully. AI-PE-RADS was grade 0 in 19 examinations (16%), grade 1 in 2 (2%), grade 2 in 3 (3%), grade 3 in 23 (19%), and grade 4 in 73 (61%). Among the 101 examinations exceeding the volume threshold, median embolic volume was 4249 mm3 (interquartile range, 2493-8774 mm3). The most proximal involved structure was the main trunk or a main pulmonary artery in 73 examinations, lobar in 23, segmental in 3, and subsegmental in 2. Grade distributions for each reader and for the consensus are given in Table 1.

**Table 1:**
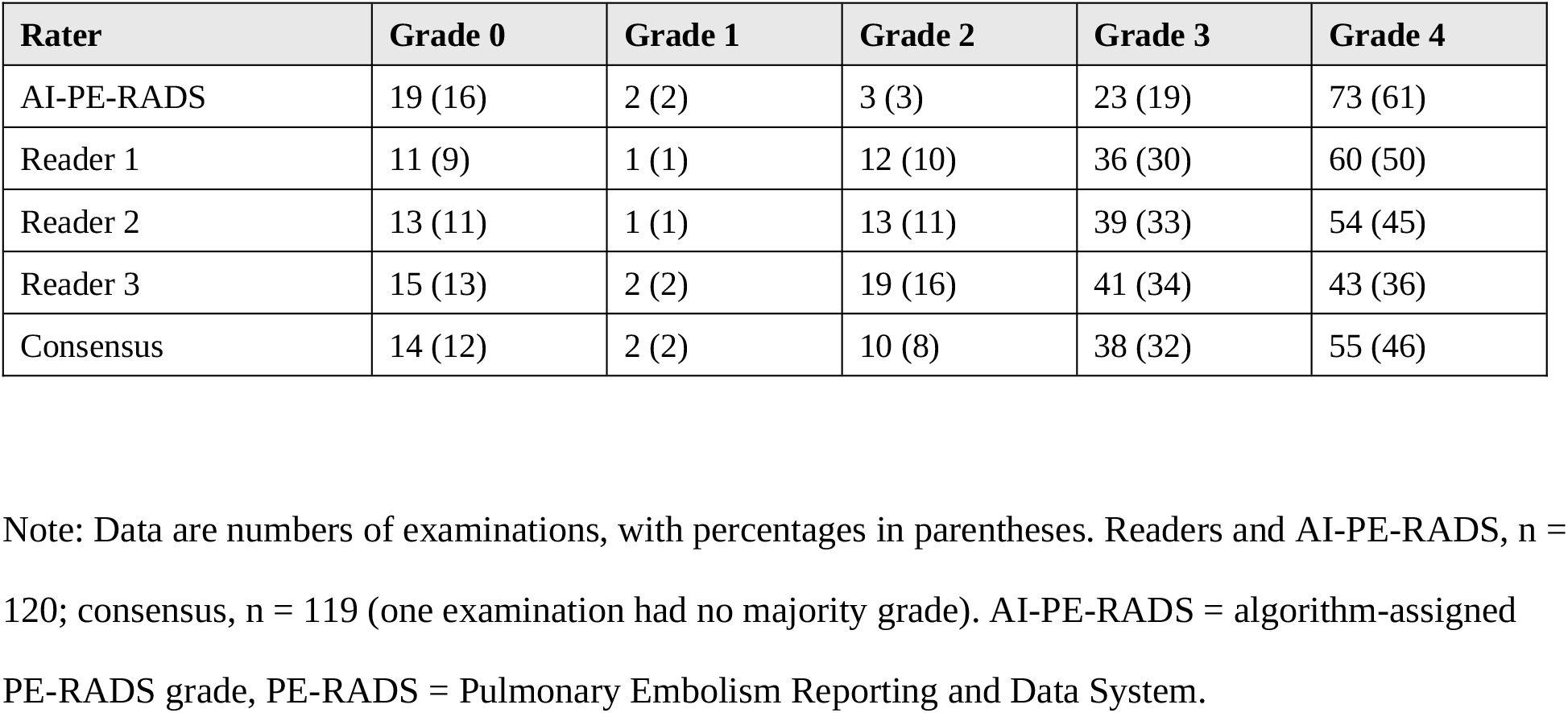
PE-RADS Grade Distribution.

| **Rater** | **Grade 0** | **Grade 1** | **Grade 2** | **Grade 3** | **Grade 4** |
| --- | --- | --- | --- | --- | --- |
| AI-PE-RADS | 19 (16) | 2 (2) | 3 (3) | 23 (19) | 73 (61) |
| Reader 1 | 11 (9) | 1 (1) | 12 (10) | 36 (30) | 60 (50) |
| Reader 2 | 13 (11) | 1 (1) | 13 (11) | 39 (33) | 54 (45) |
| Reader 3 | 15 (13) | 2 (2) | 19 (16) | 41 (34) | 43 (36) |
| Consensus | 14 (12) | 2 (2) | 10 (8) | 38 (32) | 55 (46) |

Note: Data are numbers of examinations, with percentages in parentheses. Readers and AI-PE-RADS, n = 120; consensus, n = 119 (one examination had no majority grade). AI-PE-RADS = algorithm-assigned PE-RADS grade, PE-RADS = Pulmonary Embolism Reporting and Data System.

### Agreement between AI-PE-RADS and radiologist consensus

A majority grade was available in 119 of 120 examinations. Agreement between AI-PE-RADS and consensus was 60.5% (72 of 119; kappa, 0.38; 95% CI: 0.24, 0.51) on the five-grade scale and 90.8% (108 of 119; kappa, 0.74; 95% CI: 0.59, 0.87) on the grouped scale (Fig 2, Fig 6). Of 47 discordant examinations on the five-grade scale, 40 involved adjacent grades (Fig 3, Fig 4) and 34 lay entirely within grades 3 and 4, most often consensus grade 3 against AI-PE-RADS grade 4 (24 examinations). When grading was reduced to presence or absence of embolism, AI-PE-RADS and consensus disagreed in 5 of 119 examinations (4.2%), in each case with AI-PE-RADS grade 0 where consensus indicated embolism.

**Figure 2.**
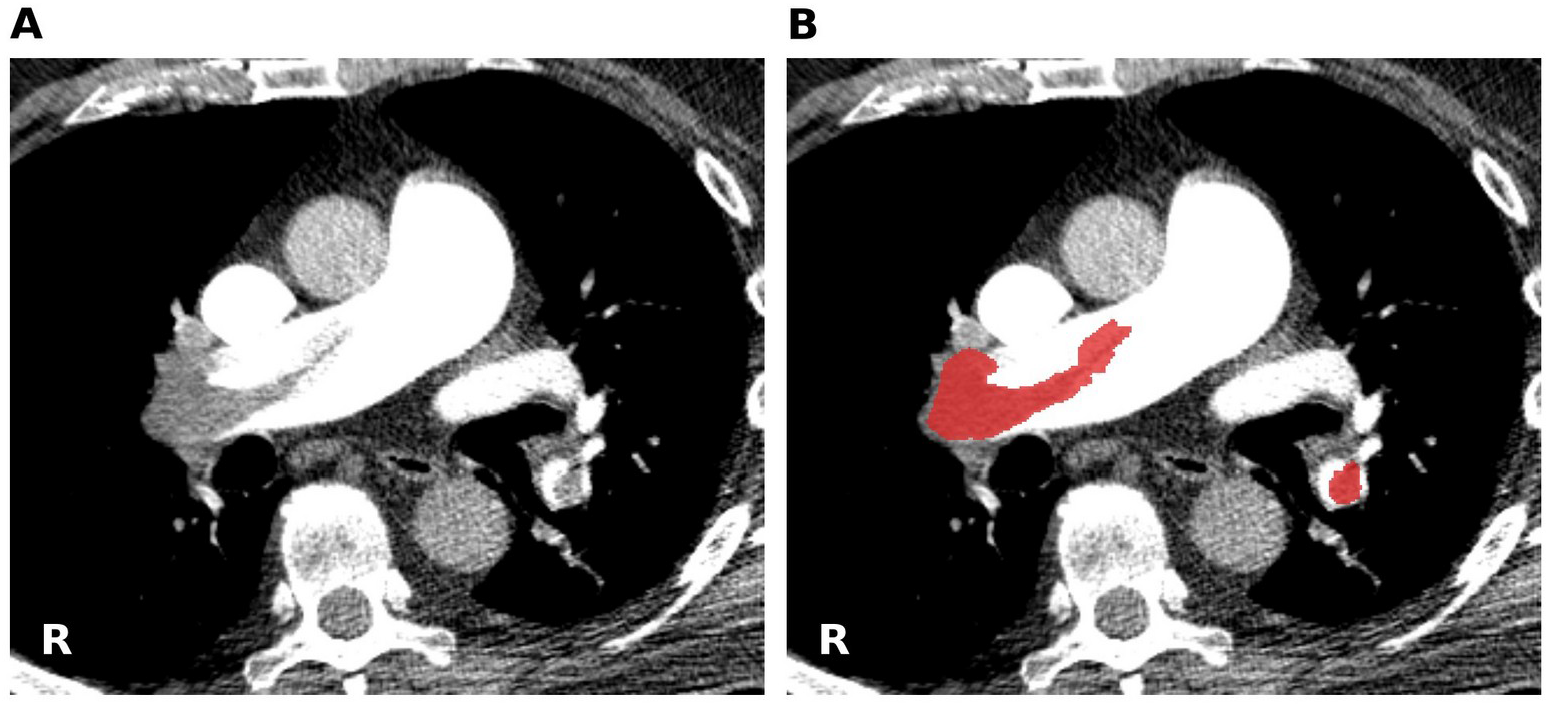
Central pulmonary embolism with full agreement between the algorithm and all three radiologists. (A) Axial CT, mediastinal window, without overlay. (B) Same section with the detected embolus overlaid in red, occupying the right main pulmonary artery. AI-PE-RADS grade 4; Readers 1, 2, and 3 each grade 4; consensus grade 4.

**Figure 3.**
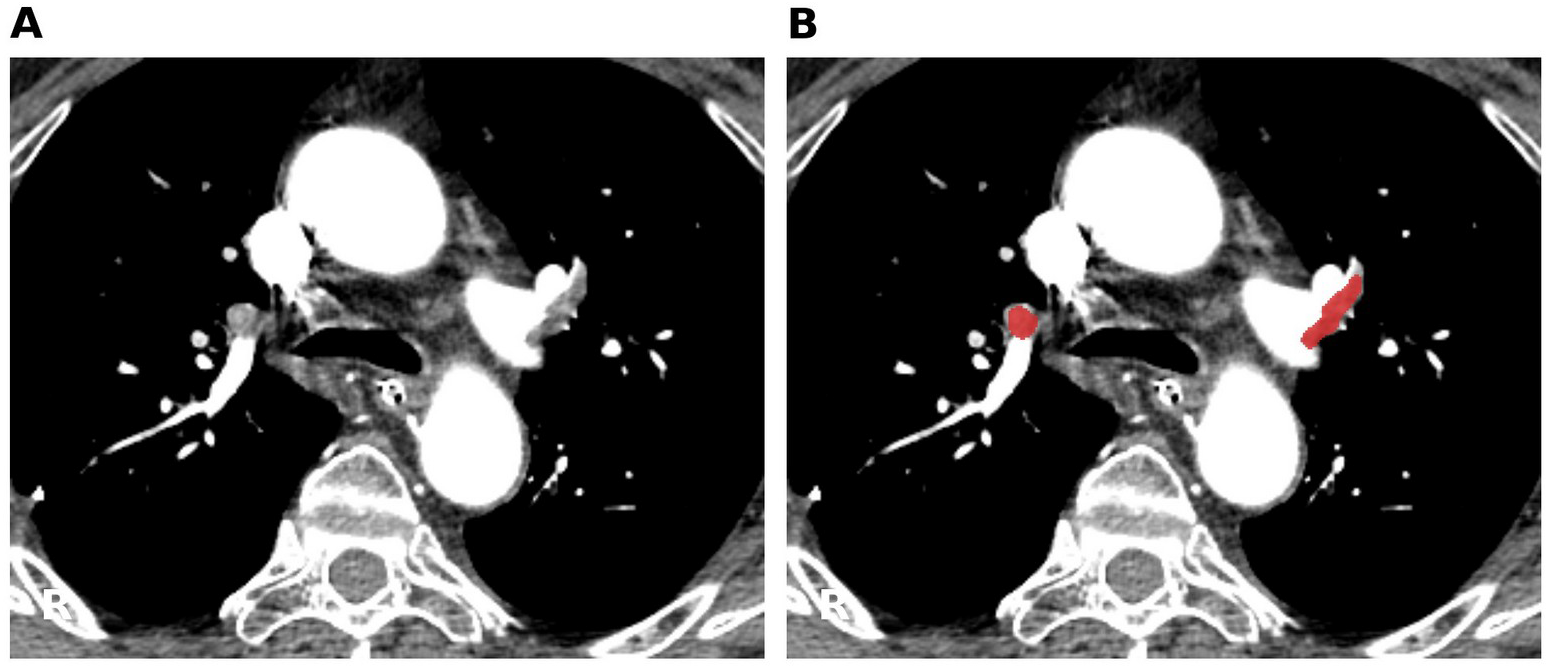
Bilateral pulmonary embolism with disagreement on the exact grade among readers and between readers and the algorithm. (A) Axial CT, mediastinal window, without overlay. (B) Same section with the detected embolus overlaid in red. The pipeline localized the embolic material to lobar branches and assigned grade 3, whereas two readers judged involvement to reach the main pulmonary artery level. AI-PE-RADS grade 3; Reader 1 grade 4; Reader 2 grade 4; Reader 3 grade 3; consensus grade 4.

**Figure 4.**
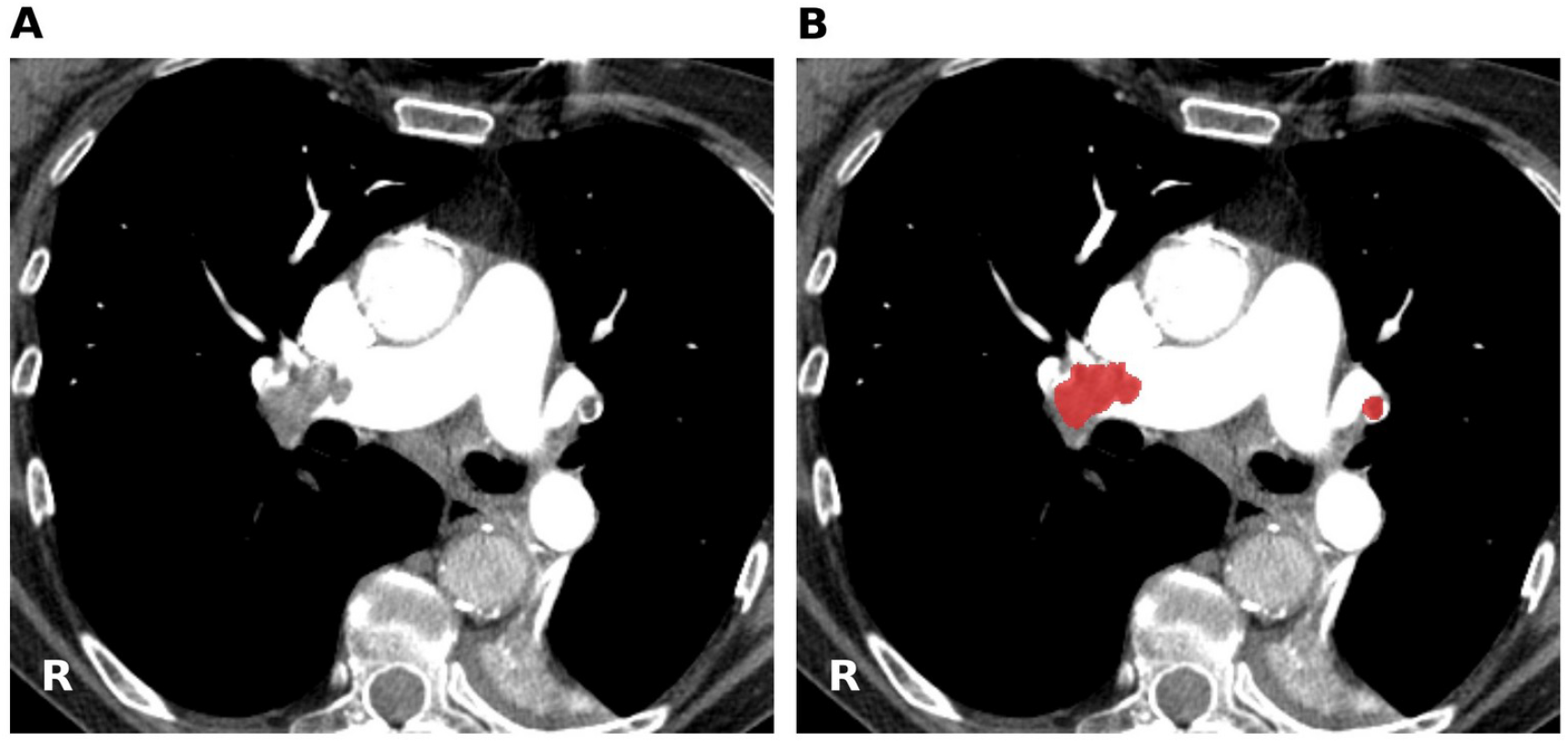
Discordant example in which the algorithm assigned a grade one level below consensus. (A) Axial CT, mediastinal window, without overlay. (B) Same section with the detected embolus overlaid in red at the right pulmonary artery bifurcation, with a small contralateral satellite embolus. The embolus was detected, but the branch carrying it was labeled lobar rather than main, yielding grade 3 where all three readers assigned grade 4. AI-PE-RADS grade 3; Readers 1, 2, and 3 each grade 4; consensus grade 4.

**Figure 5.**
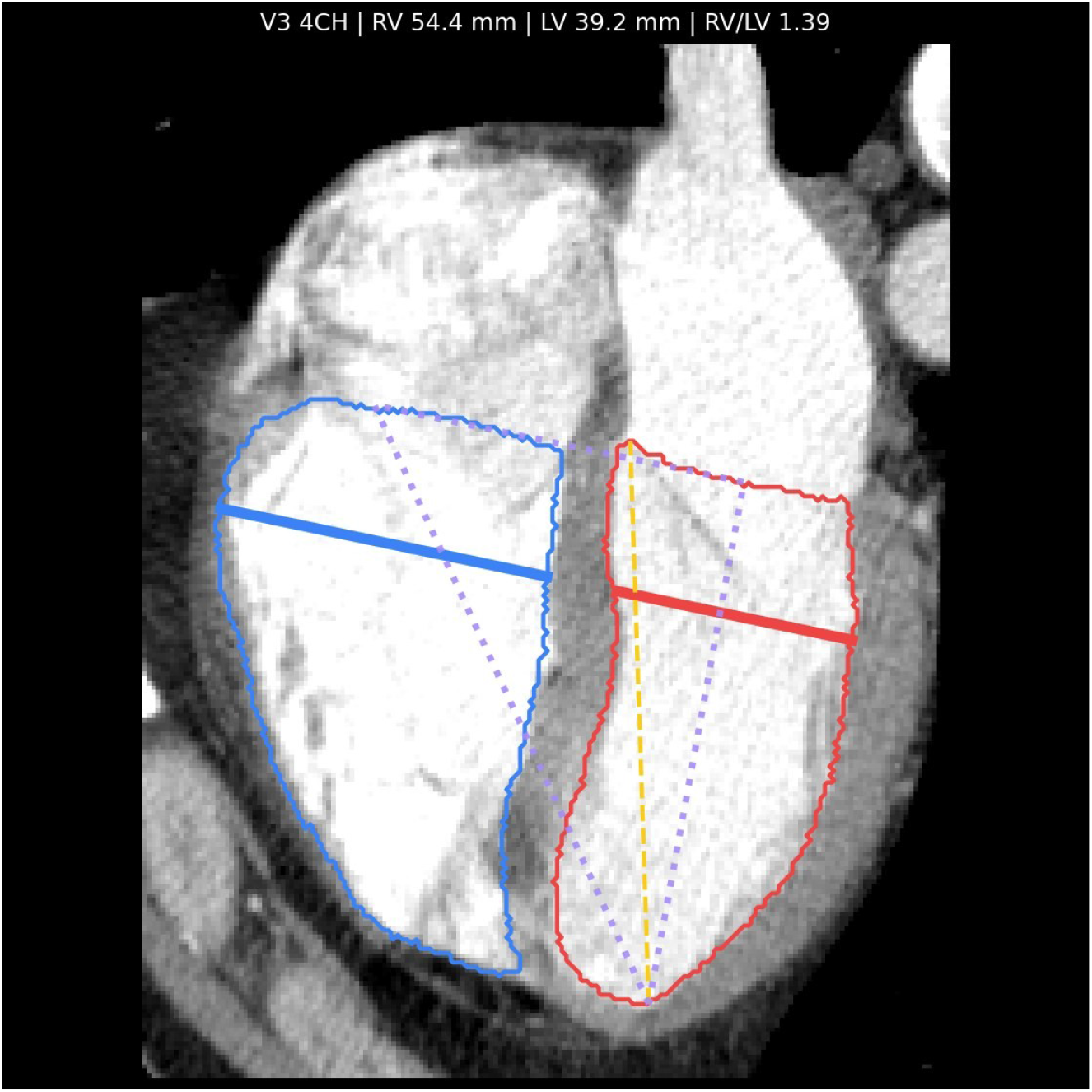
Four-chamber right ventricle-to-left ventricle (RV/LV) diameter ratio method. Reformatted four-chamber section with the right ventricle (blue outline) and left ventricle (red outline) segmented by TotalSegmentator. The measurement plane (purple dotted lines) locates the right and left ventricular internal chord diameters at 25% of the base-to-apex height, excluding the interventricular septum; the RV/LV ratio is the ratio of the two diameters. Example case: right ventricle 54.4 mm, left ventricle 39.2 mm, ratio 1.39.

**Figure 6.**
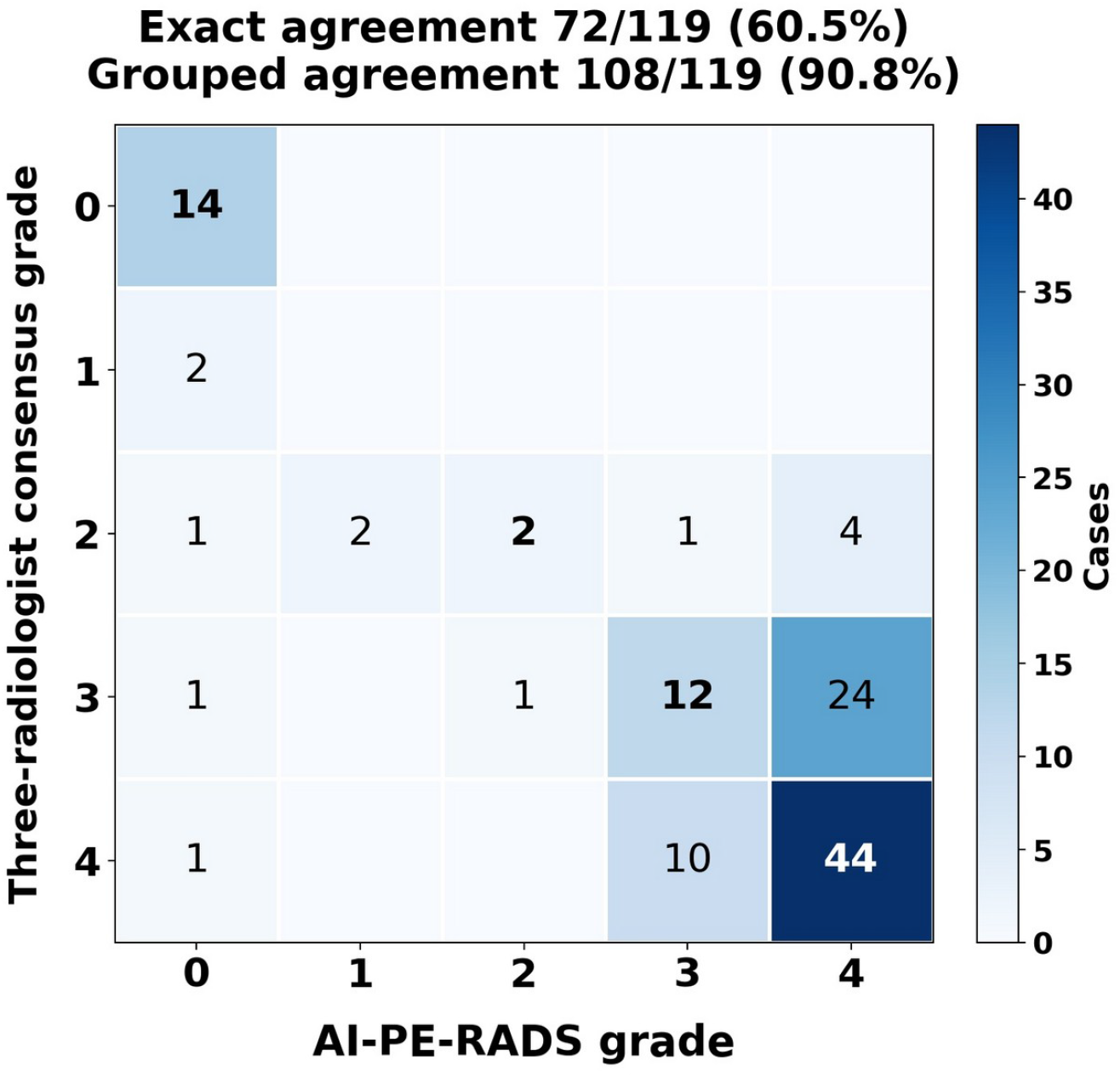
Confusion matrix of PE-RADS grade agreement between the algorithm and the three-radiologist consensus (n = 119; one examination excluded for lack of a reader majority). Rows, three-radiologist consensus grade; columns, AI-PE-RADS grade. Diagonal cells (outlined) indicate exact agreement (72 of 119, 60.5%); collapsing to the clinically grouped scheme (grade 0 versus 1-2 versus 3-4) yields agreement in 108 of 119 (90.8%).

### Agreement with individual radiologists

Pairwise agreement values are given in Table 2. On the five-grade scale, agreement between radiologists ranged from 64.2% to 79.2% (kappa, 0.48-0.68; mean kappa, 0.56), while agreement between each radiologist and AI-PE-RADS ranged from 55.8% to 58.3% (kappa, 0.32-0.36; mean kappa, 0.34). On the grouped scale the two sets converged: 84.2% to 90.8% between radiologists (kappa, 0.63-0.74; mean kappa, 0.67) and 81.7% to 90.0% between radiologists and AI-PE-RADS (kappa, 0.56-0.71; mean kappa, 0.62). Fleiss kappa among the three radiologists was 0.55 (95% CI: 0.46, 0.64) on the five-grade scale and 0.66 (95% CI: 0.53, 0.77) on the grouped scale; adding the algorithm as a fourth rater gave 0.45 (95% CI: 0.36, 0.53) and 0.64 (95% CI: 0.53, 0.74), respectively.

**Table 2:** Agreement for PE-RADS Grading.

| **Comparison** | **Agreement, five-grade (%)** | **Kappa, five-grade (95% CI)** | **Agreement, grouped (%)** | **Kappa, grouped (95% CI)** |
| --- | --- | --- | --- | --- |
| Reader 1 vs Reader 2 | 79.2 | 0.68 (0.57, 0.79) | 90.8 | 0.74 (0.60, 0.87) |
| Reader 1 vs Reader 3 | 64.2 | 0.48 (0.36, 0.60) | 85.0 | 0.63 (0.48, 0.77) |
| Reader 2 vs Reader 3 | 65.8 | 0.51 (0.38, 0.63) | 84.2 | 0.63 (0.46, 0.76) |
| Reader 1 vs AI-PE-RADS | 57.5 | 0.32 (0.17, 0.45) | 90.0 | 0.71 (0.56, 0.84) |
| Reader 2 vs AI-PE-RADS | 58.3 | 0.35 (0.22, 0.48) | 85.8 | 0.60 (0.44, 0.75) |
| Reader 3 vs AI-PE-RADS | 55.8 | 0.36 (0.24, 0.48) | 81.7 | 0.56 (0.40, 0.70) |
| AI-PE-RADS vs Consensus | 60.5 | 0.38 (0.24, 0.51) | 90.8 | 0.74 (0.59, 0.87) |
| Three readers (Fleiss) | ... | 0.55 (0.46, 0.64) | ... | 0.66 (0.53, 0.77) |
| Three readers plus AI-PE-RADS (Fleiss) | ... | 0.45 (0.36, 0.53) | ... | 0.64 (0.53, 0.74) |

Note: Grouped scale is grade 0 versus grades 1-2 versus grades 3-4. Reader-reader and reader-AI-PE-RADS comparisons, n = 120; comparisons involving consensus, n = 119. Confidence intervals were obtained by bootstrap resampling with 5000 replicates. AI-PE-RADS = algorithm-assigned PE-RADS grade, CI = confidence interval, PE-RADS = Pulmonary Embolism Reporting and Data System.

### Segmentation quality

At least one radiologist flagged embolus segmentation as incomplete in 32 of 120 examinations (27%), with marked differences between readers (2 of 120 [2%], 11 of 120 [9%], and 24 of 120 [20%]). No examination was flagged by all three readers and only 5 by two, indicating reader-specific thresholds rather than a consistently identified subset of poorly segmented cases.

### RV/LV ratio

All three radiologists rated RV/LV plausibility in all 120 examinations. Pairwise agreement on the favorable-versus-incorrect dichotomy was 75.8% (kappa, 0.03), 59.2% (kappa, 0.14), and 56.7% (kappa, 0.04), and Fleiss kappa was −0.00 (95% CI: −0.09, 0.09), indistinguishable from chance. Rating distributions diverged markedly: one reader rated 2 of 120 measurements incorrect, another 29, and the third 54. Results were similar on the three-level scale (Fleiss kappa, −0.01; 95% CI: −0.08, 0.05).

In the external comparison, 2119 of 2211 labeled examinations (95.8%) had both a pipeline ratio and the dataset label. Agreement was 74.0% (95% CI: 72.1, 75.8) with a kappa of 0.49 (95% CI: 0.45, 0.52). Disagreement was directional: the pipeline indicated a ratio of at least 1 in 416 examinations labeled below 1, against 135 in the opposite direction, approximately 3.1 to 1. The pipeline classified 1187 of 2119 examinations (56.0%) as having a ratio of at least 1, compared with 906 (42.8%) by the dataset label.

## Discussion

AI-PE-RADS grades agreed substantially with a three-radiologist consensus on the clinically grouped scale and, as a fourth reader, reached grouped-scale agreement comparable to that observed among the radiologists themselves. Agreement was lower on the ungrouped five-grade scale, but nearly all residual disagreement involved adjacent grades within the severe range, and disagreement about whether embolism was present at all was rare.

This pattern suggests the pipeline reproduces the clinically consequential distinction, between absent, peripheral, and central or lobar disease, more reliably than finer adjacent-grade distinctions. This grouping reflects PE-RADS management pathways (no treatment, anticoagulation, or reperfusion consideration), so grouped-scale agreement approximates management-relevant agreement (3). The finer, ungrouped distinction was the least reproducible among radiologists, consistent with the boundary between lobar and more proximal involvement depending on judgments about branch identity that are graded rather than categorical.

The RV/LV findings illustrate a different problem. Radiologists showed no agreement when asked whether an automated four-chamber measurement was plausible, whether rated on the three-level scale or dichotomized as favorable versus incorrect, with distributions so divergent that readers were applying different internal thresholds. That the same measurement agreed moderately with an independent binary label across more than 2000 examinations suggests the absence of reader agreement reflects a poorly operationalized subjective judgment rather than measurement failure. This does not exculpate the algorithm: the external comparison also revealed a systematic tendency to overestimate ventricular strain by approximately three to one, a limitation. CTPA is not electrocardiographically gated, and the choice of measurement plane, axial versus reformatted four-chamber, is known to affect the RV/LV ratio (17).

This study has several limitations. No independent reference standard for PE-RADS grade exists, so all comparisons are of agreement rather than accuracy; detection accuracy was established separately in a dual-cohort study (14). The consensus derives from the same three readers whose agreement is characterized. The sample is modest and enriched for central disease, so the grade distribution does not reflect clinical prevalence. Readers viewed the algorithm segmentation while grading, which may have anchored their assessments. As noted in Methods, the training and evaluation datasets differ in origin. Finally, the RV/LV design asked readers only about anatomic plausibility; a quantitative reader measurement would have permitted a stronger comparison.

In this preliminary evaluation, automated PE-RADS grading performed comparably to a fourth reader on the grouped scale, while automated RV/LV measurement showed a systematic bias requiring correction before clinical evaluation. The pipeline is a research tool and is not intended for clinical use.

## Data Availability

All data produced are available online at https://github.com/eziolanza/perads-net

## Abbreviations

AI-PE-RADS: algorithm-assigned PE-RADS grade
CI: confidence interval
CTPA: CT pulmonary angiography
LV: left ventricle
PE: pulmonary embolism
PE-RADS: Pulmonary Embolism Reporting and Data System
RV: right ventricle

## Declarations

## Ethics approval

The evaluation data are public and fully de-identified, and their secondary analysis required no ethics committee review. The development cohort was collected under an institutional policy whereby retrospective studies not affecting clinical care proceed without ethics submission.

## Consent to participate

Waiver of written informed consent applied to the development cohort under the institutional policy described above. The evaluation dataset is publicly available and de-identified at source.

## Data and code availability

The complete PERADS.net pipeline, including inference code and trained model weights, is publicly available at https://github.com/eziolanza/perads-net (release v0.2.0), under a CC BY-NC 4.0 license. The per-examination data underlying the dataset-wide right-to-left ventricular ratio comparison are provided as a supplementary file (atlas_rsna_rvlv_compact.csv). The imaging data are the publicly available RSNA Pulmonary Embolism CT dataset (references 8 and 9).

## Author contributions

E.L.: conceptualization, data curation, investigation, methodology, project administration, software, validation, writing - original draft, writing - review and editing. F.C.: validation, writing - review and editing. C.L.: validation, writing - review and editing. F.D.: validation. R.L.: conceptualization, resources, software. A.L.: supervision, validation, writing - review and editing.

