## Supplemental files for "PERADS.net: Automated PE-RADS Grading with Named Anatomic Localization and Right-to-Left Ventricular Ratio Measurement on CT Pulmonary Angiography"

### Supplementary Material

### PERADS.net: Code, Model, and Data Availability

#### S1. Overview

PERADS.net is released as an open, reference implementation of the pipeline described in the main manuscript. The complete source code, installation instructions, and the trained nnU-Net model weights used for embolus segmentation are publicly available on GitHub at:

`https://github.com/eziolanza/perads-net`

The repository is intended to let readers reproduce the method end to end, from a single raw, uncropped chest CT pulmonary angiogram (NIfTI format) to a PE-RADS grade, named anatomic site, embolic burden, and (optionally) an RV/LV diameter ratio, without access to the training data or any part of the internal research infrastructure used to develop it.

#### S2. Repository contents

The repository contains:

- perads_net_pipeline.py - the single-case, end-to-end pipeline (TotalSegmentator, crop, nnU-Net embolus segmentation, named-artery PE-RADS grading, optional RV/LV ratio).
- batch_perads_net_pipeline.py - a thin multi-case wrapper around the single-case pipeline (manifest CSV or directory input, configurable worker count).
- rvlv_v3_four_chamber.py - the four-chamber RV/LV ratio measurement module.
- requirements.txt - pinned Python dependencies.
- README.md - installation and usage instructions (summarized in this document).
- LICENSE - the code license (CC BY-NC 4.0).

The trained nnU-Net model weights (five-fold three-dimensional full-resolution ensemble, approximately 1.1 GB compressed) are distributed separately as a GitHub Release asset rather than committed to the repository itself, following standard practice for binary model artifacts.

### S3. Installation

```
git clone https://github.com/eziolanza/perads-net.git
cd perads-net
pip install -r requirements.txt
```

An NVIDIA GPU with CUDA is required, together with TotalSegmentator and nnUNetv2_predict_from_modelfolder available on the system.

The trained model bundle is downloaded from the repository's GitHub Releases page:

```
wget
https://github.com/eziolanza/perads-net/releases/download/v0.2.0/PERADS
.net-model-v0.2.0.tar.zst
tar --use-compress-program=unzstd -xf PERADS.net-model-v0.2.0.tar.zst
```

This extracts a nnUNetTrainer__nnUNetPlans__3d_fullres/ folder (dataset.json, plans.json, fold_0 through fold_4). The pipeline is pointed at it either with the --model command-line argument or the PERADS_NET_MODEL environment variable.

### S4. Usage

#### S4.1 Full pipeline (PE-RADS grade and RV/LV ratio)

```
python3 perads_net_pipeline.py \
  --input /absolute/path/to/raw_ct.nii.gz \
  --output /absolute/path/to/output_case \
  --device cuda
```

#### S4.2 PE-RADS grade only (no RV/LV, no additional license required)

The RV/LV step requires TotalSegmentator's heartchambers_highres task (see Section S5). Users who only need the PE-RADS grade and anatomic site, and who do not have, or do not want to request, that TotalSegmentator license, can skip this step entirely with the --skip-rvlv flag. This runs only the license-free segmentation tasks (pulmonary arteries, lung lobes) and the embolus segmentation and grading steps; the three RV/LV-related fields in the output are left empty.

```
python3 perads_net_pipeline.py \
  --input /absolute/path/to/raw_ct.nii.gz \
  --output /absolute/path/to/output_case \
  --device cuda --skip-rvlv
```

#### S4.3 Batch processing

batch_perads_net_pipeline.py accepts either a manifest CSV (a ct_path column, and optionally a case_id column) or a directory to scan recursively for raw CT NIfTI files, and runs the single-case pipeline across a configurable number of worker threads. It accepts the same --skip-rvlv flag.

```
python3 batch_perads_net_pipeline.py \
  --manifest cases.csv \
  --output-root /absolute/path/to/batch_output \
  --workers 4 --device cuda
```

Both the single-case and batch entry points re-use per-case TotalSegmentator and nnU-Net outputs already present on disk (idempotent execution), so re-running after an interruption, or with --overwrite, only recomputes the missing steps.

### S5. TotalSegmentator licensing for the RV/LV step

The RV/LV ratio calculation depends on TotalSegmentator's heartchambers_highres task for right and left atrium and ventricle segmentation. Unlike the tasks used for the PE-RADS grading pathway (lung_vessels, lobe segmentation via the total task), heartchambers_highres is not covered by TotalSegmentator's default, license-free tasks: it requires requesting a separate, free license directly from the TotalSegmentator authors. Instructions for requesting this license are provided in the official TotalSegmentator repository:

```
https://github.com/wasserth/TotalSegmentator
```

Readers who wish to reproduce only the PE-RADS grading results reported in the manuscript, without the RV/LV ratio, do not need to request this license: passing --skip-rvlv (Section S4.2) avoids invoking heartchambers_highres entirely.

### S6. Output format

For each processed case, the pipeline writes a case subdirectory containing intermediate TotalSegmentator and nnU-Net outputs, a structure_labelmap.nii.gz (the named-artery classification volume), and a results/result.json with the full per-case result. One summary row per case is also appended to a run-level CSV with the following fields:

- case_id
- perads_grade - PE-RADS grade, 0 to 4
- anatomic_level - named anatomic level of the most proximal embolus
- most_proximal_structures - named artery segment or segments at that level
- embolic_voxels, bcv_mm3 - predicted embolic burden (voxel count and volume, mm3)
- classification_reason
- rv_lv_ratio, rv_diameter_mm, lv_diameter_mm - empty when --skip-rvlv is used
- num_branches, duration_sec

### S7. Versioning and reproducibility

The GitHub Release tagged v0.2.0 corresponds to the exact pipeline version and trained model weights used to generate the results reported in this manuscript, including the 70 mm3 minimum embolus volume threshold and the removal of the arterial hard-mask from embolus grading described in Materials and Methods. Cloning the repository at this tag, together with the v0.2.0 model release asset, reproduces the exact configuration evaluated in this study.

### S8. Supplementary data file: atlas_rsna_rvlv_compact.csv

The dataset-wide comparison of the automated right-to-left ventricular (RV/LV) diameter ratio against the RSNA reference label, reported in the Results section of the main manuscript, is provided in full as a single comma-separated file, atlas_rsna_rvlv_compact.csv. The file allows that analysis to be recomputed independently, without running the pipeline and without access to the source images.

#### S8.1 Provenance

Each row corresponds to one CT pulmonary angiography examination of the publicly available RSNA Pulmonary Embolism CT dataset (RSPECT) training split, as distributed by the Radiological Society of North America. The reference column is taken unchanged from the RSNA study-level annotations; the algorithm column is the output of PERADS.net v0.2.0 applied to that examination. No image data, patient identifiers, dates, or institution labels are included, and the case identifier is the de-identified study identifier already published as part of the RSNA dataset, so the file introduces no additional re-identification risk.

#### S8.2 Structure

The file contains 7279 rows (one per examination, all identifiers unique) and three columns:

- case_id - the de-identified RSNA study identifier (12-character hexadecimal string). Populated for all 7279 rows.
- rv_lv_rsna - the RSNA study-level RV/LV reference label, recorded as the literal strings "<1" or ">=1". Populated for 2211 rows (940 ">=1" and 1271 "<1"). The field is empty for the remaining 5068 examinations because RSNA assigns this label only to examinations reported as positive for pulmonary embolism.

- rv_lv_algorithm - the continuous four-chamber RV/LV diameter ratio produced by PERADS.net, range 0.34 to 2.43. Populated for 7024 rows; empty where the pipeline did not return a ratio.

Both fields are populated in 2119 examinations, that is 95.8% of the 2211 labeled examinations. These 2119 examinations constitute the analysis set for the external RV/LV comparison reported in the manuscript. The file is deliberately restricted to these three columns: it carries the reference label and the algorithm output and no derived or intermediate quantity, so that every reported value can be recomputed from it and audited directly.

#### S8.3 Recomputing the reported results

Dichotomizing rv_lv_algorithm at a threshold of 1.0 and cross-tabulating it against rv_lv_rsna, restricted to the rows in which both fields are populated, reproduces every value reported in the manuscript: 74.0% raw agreement, Cohen kappa 0.49, 416 examinations classified by the pipeline as having a ratio of at least 1 against an RSNA label below 1 versus 135 in the opposite direction, and 1187 examinations (56.0%) classified by the pipeline as having a ratio of at least 1 against 906 (42.8%) by the RSNA label. The following is sufficient to reproduce these values:

```
import pandas as pd
from sklearn.metrics import cohen_kappa_score

df = pd.read_csv("atlas_rsna_rvlv_compact.csv")
df = df.dropna(subset=["rv_lv_rsna", "rv_lv_algorithm"])

reference = (df["rv_lv_rsna"] == ">=1").astype(int)
predicted = (df["rv_lv_algorithm"] >= 1.0).astype(int)

print(len(df))
print((reference == predicted).mean())
print(cohen_kappa_score(reference, predicted))
```

### S9. Disclaimer and license

This is a research pipeline. It has not been validated for clinical use and requires expert visual review of every case; it must not be used to guide patient management. The code is released under a CC BY-NC 4.0 license (non-commercial use with attribution).
